# Long-Term Subcortical Electrophysiological Recordings Link Heightened Interhemispheric Subthalamic Beta Synchrony to Progression of Bradykinesia in Parkinson’s Disease

**DOI:** 10.1101/2022.09.13.22279778

**Authors:** Kevin B. Wilkins, Yasmine M. Kehnemouyi, Matthew N. Petrucci, Ross W. Anderson, Jordan E. Parker, Megan H. Trager, Raumin S. Neuville, Mandy M. Koop, Anca Velisar, Zack Blumenfeld, Emma J. Quinn, Helen M. Bronte-Stewart

## Abstract

Bradykinesia is the major cardinal motor sign of Parkinson’s disease (PD), but its neural underpinnings are unclear. Although impairment in PD has been linked to heightened synchrony within the beta band (13-30 Hz) in the subthalamic nucleus (STN), bradykinesia appears to be the manifestation of a network-level dysfunction including the basal ganglia, motor cortex, and possibly cerebellum. The goal of this study was to examine whether changes in bradykinesia over time following long-term STN deep brain stimulation (DBS) are linked to local STN beta dynamics or a wider bilateral network dysfunction. Twenty-one individuals with Parkinson’s disease chronically implanted with sensing neurostimulators (Activa® PC+S, Medtronic, PLC) in the STN participated in a longitudinal ‘washout’ therapy study every three to six months for an average of three years. At each visit, participants were withdrawn from medication (12/24/48 hours) and had DBS turned off (>60 minutes) prior to completing a repetitive wrist-flexion extension task, a validated quantitative assessment of bradykinesia. Synchronized local field potentials and kinematic data were recorded. Local STN beta dynamics were investigated via STN beta power and burst duration, while interhemispheric beta synchrony was assessed with STN beta coherence. Higher beta power and interhemispheric STN beta coherence, but not burst duration, were significantly associated with worse bradykinesia. Bradykinesia was found to worsen off therapy over time. Interhemispheric STN beta coherence also increased over time, whereas beta power and burst duration remained stable. The observed percent change in bradykinesia was related to the percent change in interhemispheric beta coherence, with greater increases in synchrony associated with further worsening of bradykinesia. Together, these findings implicate interhemispheric beta synchrony as a neural correlate of the progression of bradykinesia following chronic STN DBS. This could imply the existence of a pathological bilateral network contributing to bradykinesia in PD.

## Introduction

Bradykinesia, defined as slowness of movement, is one of the cardinal motor signs of Parkinson’s disease (PD). Both dopaminergic medication and either subthalamic nucleus (STN) or globus pallidus interna (GPi) deep brain stimulation (DBS) substantially improve aspects of bradykinesia. However, the progressive nature of PD often leads to its worsening over time despite optimal treatment. The underlying neural mechanisms of bradykinesia and its potential longitudinal progression are still unclear.

The basal ganglia, primary motor cortex (M1), and cerebellum have all been implicated in contributing to bradykinesia in PD suggesting a network-level dysfunction^1^. Recordings of local field potentials (LFPs) in the STN have revealed exaggerated neuronal oscillations and synchrony in the beta frequency band (13-30 Hz) as a marker of pathophysiology of Parkinson’s disease, including bradykinesia^2–4^. Similarly, reductions in beta power and burst duration with medication and/or DBS are associated with its improvement^5–7^. This heightened synchrony does not exist solely in the basal ganglia, but instead appears to propagate throughout the basal ganglia-thalamo-cortical loop, as evident by simultaneous bilateral STN and subcortical-cortical recordings^8–14^. Entraining beta within this circuit via transcranial alternate current stimulation over the primary motor cortex has been shown to worsen bradykinesia^15^. Furthermore, the potential network dysfunction contributing to bradykinesia may not be limited only to the contralateral side of the basal ganglia-thalamo-cortical loop. Unilateral DBS induces ipsilateral improvement in bradykinesia, implicating a potential bilateral contribution^16–20^. In support of this possibility, heightened bilateral beta synchrony between STNs has been shown despite no known direct anatomical connections between these structures^8,10,21^.

The advent of chronically implanted sensing neurostimulators has enabled the ability to track subcortical basal ganglia dynamics longitudinally alongside potential disease progression. Interestingly, the largest multi-year longitudinal LFP study in PD to date found that STN beta bursts and beta power measured at rest were stable over time and no worsening of overall clinical motor impairment was observed^22^. However, when examining symptom-specific changes over time, there is evidence that bradykinesia and axial symptoms may continue to worsen with chronic STN DBS^23,24^, compared to tremor, which improves over time^25,26^. Given the apparent stability of STN beta power and burst duration over time, it is possible that worsening of bradykinesia may be due to a wider network dysfunction.

The goal of this study was to examine whether changes in bradykinesia over time following long-term STN DBS are linked to local STN beta dynamics or a wider network dysfunction. We combined synchronized bilateral STN neural activity and quantitative kinematic data alongside a therapy washout protocol (i.e., after temporary withdrawal of medication and STN DBS) every three to six months for up to seven years (average of three years) to track progression of bradykinesia and its neural correlates. We investigated whether either local STN beta dynamics or interhemispheric STN beta synchrony related to changes in bradykinesia over time. We hypothesized that worsening of bradykinesia over time would be associated with increased interhemispheric STN beta synchrony, but not changes in local STN beta dynamics.

## Materials and Methods

### Human Subjects

Twenty-one participants (16 male, 5 female) with clinically established Parkinson’s disease (Table 1) underwent bilateral implantation of DBS leads (model 3389, Medtronic, PLC) in the sensorimotor region of the subthalamic nucleus (STN) using a standard functional frameless stereotactic technique and multi-pass microelectrode recording. Dorsal and ventral borders of each STN were determined using microelectrode recording, and the base of electrode zero was placed at the ventral border of the STN. The two leads were connected to the implanted investigative neurostimulator (Activa® PC+S, Medtronic, PLC). The preoperative selection criteria and surgical technique have been previously described^27^. All experimental testing was done in the off-medication state, which entailed stopping long-acting dopamine agonists at least 48 hours, dopamine agonists and controlled release carbidopa/levodopa at least 24 hours, and short acting medication at least 12 hours before testing. All participants gave written consent to participate in the study, which was approved by the Food and Drug Administration (FDA) with an Investigational Device Exemption (IDE) and by the Stanford University Institutional Review Board (IRB).

**Table 1.** Participant Demographics

| ID | Age | Sex | Disease Duration (Years) | Pre-Op UPDRS III (off meds) | Pre-Op UPDRS III (on meds) | IP MDS-UPDRS III (off therapy) | # of Visits | Length of Time (Months) |
| --- | --- | --- | --- | --- | --- | --- | --- | --- |
| 1 | 71-75 | M | 9.9 | 54 | 41 | 60 | 4 | 27 |
| 2 | 51-55 | M | 4.9 | 24 | 8 | 20 | 12 | 63 |
| 3 | 61-65 | M | 4.0 | 35 | 26 | 36 | 10 | 69 |
| 4 | 61-65 | M | 7.2 | 29 | 16 | 38 | 7 | 49 |
| 5 | 56-60 | M | 7.3 | 39 | 23 | 43 | 11 | 63 |
| 6 | 41-45 | M | 7.5 | 58 | 12 | 51 | 4 | 12 |
| 7 | 66-70 | M | 10.6 | 41 | 28 | 43 | 3 | 6 |
| 8 | 51-55 | M | 7.6 | 52 | 29 | 61 | 11 | 59 |
| 9 | 71-75 | M | 9.9 | 47 | 23 | 45 | 8 | 29 |
| 10 | 56-60 | F | 11.0 | 33 | 6 | 42 | 9 | 37 |
| 11 | 51-55 | M | 11.0 | 56 | 20 | 60 | 1 | 3 |
| 12 | 31-35 | M | 8.1 | 59 | 22 | 60 | 8 | 30 |
| 13 | 56-60 | M | 6.1 | 62 | 23 | 56 | 3 | 12 |
| 14 | 66-70 | F | 7.1 | 44 | 25 | 12 | 13 | 71 |
| 15 | 46-50 | F | 11.2 | 50 | 26 | 51 | 8 | 30 |
| 16 | 61-65 | M | 17.3 | 42 | 24 | 44 | 6 | 18 |
| 17 | 51-55 | F | 12.5 | 34 | 6 | 22 | 10 | 61 |
| 18 | 66-70 | M | 8.6 | 51 | 10 | 33 | 4 | 31 |
| 19 | 56-60 | F | 13.6 | 38 | 18 | 35 | 10 | 57 |
| 20 | 51-55 | M | 12.3 | 18 | 4 | 21 | 6 | 18 |
| 21 | 46-50 | M | 11.2 | 38 | 31 | 48 | 7 | 25 |
| Avg ± SD | 57.8 ± 9.6 | 16 M, 5 F | 9.5 ± 3.1 | 43.1 ± 11.9 | 20.1 ± 9.6 | 42.4 ± 15.2 | 7.4 ± 3.3 | 36.7 ± 22.0 |

### Experimental Protocol

Experiments were conducted off all therapy every three to six months after initial programming of DBS for up to seven years. Prior to data collection, STN DBS was turned off for 60-75 minutes, which we have shown to be enough time to wash out the effect of DBS on STN LFPs^28^. Participants then performed a seated repetitive wrist flexion-extension (rWFE) task, which we have validated as a measure of bradykinesia in PD^29^. Participants were instructed to remain seated and as still as possible with their eyes open during a 30 second rest period. After a ‘Go’ command, participants flexed the forearm so that the elbow was angled at 90° and then flexed and extended the hand at the wrist joint as quickly as possible and stopped only when instructed. Participants completed the task for 30 seconds. LFPs from bilateral STNs were simultaneously recorded.

### Data Acquisition and Analysis Kinematic Data Acquisition

Movement was measured using solid-state gyroscopic wearable sensors (Motus Bioengineering, Inc.) attached to the dorsum of each hand. Data was sampled at 1 kHz. The experiment was monitored by continuous video that was synchronized to the kinematic signals.

### LFP Data Acquisition

STN LFPs were recorded from electrode contact pair 0-2 or 1-3 on the DBS leads using the Activa® PC+S system. The chosen contact pair was held constant across visits over time. LFPs were high pass filtered at 0.5 Hz, low pass filtered at 100 Hz within the device, and sampled at 422 Hz. Uncompressed neural data were recorded on to the Activa® PC+S system and then extracted via telemetry using the Activa® PC+S tablet programmer.

### Synchronization of local field potential and kinematic data

The kinematic and LFP signals were acquired concurrently, using a data acquisition interface (Power1401) and Spike software (version 2.7, Cambridge Electronic Design, Ltd., Cambridge, England). The synchronization of neural and kinematic recordings, using internal and external instrumentation, respectively, was achieved by administering a few seconds of 20 Hz/1.5 V neurostimulation through one of the DBS leads. The signal artifact was detected concurrently by the implanted system and Spike software, the latter system recording the EEG stimulation artifact using surface electrodes attached to the skin (one on the forehead, one on the collarbone, and one above the implanted neurostimulator), which were recorded at 1 kHz. The files were then co-registered in MATLAB during offline analysis.

### Kinematic Data Analysis

Angular velocity data were low-pass filtered using a zero-lag fourth order Butterworth filter with a 4 Hz cut-off frequency. Root mean square velocity (V_rms_) was calculated across the trial.

### Neural Data Analysis

Estimated power spectral density was calculated using Welch’s method with a 1 second sliding Hanning window with 50% overlap on the 30 seconds of movement data. Power was calculated across the whole beta band (13-30 Hz) and normalized by dividing these values by the mean power across the 45-65 Hz band^30^.

Beta bursts were calculated using a previously validated method across the whole beta band^31^. Briefly, this process involved filtering the raw LFP with a zero-phase 8^th^ order Butterworth bandpass filter, squaring the signal, and creating an amplitude envelope. A baseline was used for thresholding based on 4 times the median power of the troughs of the LFP within the 45 to 65 Hz band. Burst durations were calculated as the interval between successive crossings of the envelope of the band of interest over the baseline.

Synchronization between STNs was evaluated through magnitude-squared wavelet coherence using the following equation:

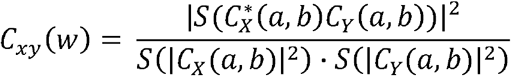

Where C_x_(a,b) and C_y_(a,b) represent the continuous wavelet transforms of signals *x* and *y* at scales of *a* and positions *b*. The superscript * is the complex conjugate and *S* is a smoothing operator in time and scale. Wavelets were used due to lack of assumption regarding stationarity of the neural signal during movement. The average coherence within the beta band (13-30 Hz) across the trial was calculated.

### Statistical Analysis

Statistical analyses were run in R (version 3.6.0, R Foundation for Statistical Computing, University of Auckland, New Zealand) and MATLAB (version 9.9, MathWorks, Natick MA). Behavioral and neural metrics were first converted into z-scores. To assess whether there was a relationship between any neural metric and behavior during the rWFE task, a linear mixed effects model was used with a fixed effect of the neural metric of interest and a random intercept for subject and a dependent variable of V_rms_. This model was run for normalized beta power, beta burst duration, and beta coherence. To assess changes over time in either behavior or neural metrics, a linear mixed effects model was used with a fixed effect of time, a random slope of time, and a random intercept for subject. This model was run for V_rms_, normalized beta power, beta burst duration, and beta coherence.

Lastly, to determine any relationship between change in neural metrics over time and change in behavior over time the following procedure was used: 1. The percent change in a given metric from one visit to the next was calculated; 2. The percent change was transformed into a percent change per month by dividing the percent change by the duration of time that elapsed between subsequent visits; 3. A linear mixed effects model was used with a fixed effect of time and percent change per month of a given neural metric, a random slope of time, and a random intercept for subjects. The dependent variable was the percent change per month of V_rms_. This model was run for normalized beta power, beta burst duration, and beta coherence. Standardized beta coefficients and subsequent 95% confidence intervals are reported for all linear mixed effect models. All reported p-values were corrected with a Bonferroni correction for the number of models tested. Significance was set at *p* < 0.05 Bonferroni corrected.

## Results

### Demographic Characteristics

Demographic information of the twenty-one individuals with PD is shown in Table 1. The participants had a mean MDS-UPDRS III of 42.4 ± 15.2 off therapy at initial programming and a disease duration of 9.5 ± 3.1 years. Participants completed on average 7.4 ± 3.3 visits that spanned 36.7 ± 22.0 months. Variability in number of visits was due to lack of re-implantation of the experimental neurostimulator (PC+S), later enrollment in the study, or inability to tolerate off therapy testing.

### Increased STN Beta Coherence is Associated with Worse Bradykinesia

Higher beta coherence between STNs was significantly associated with lower (i.e., worse) V_rms_ during rWFE (β = -0.24 [-0.34 -0.13], t = 4.25, *p* = 8.73e-5; Figure 1). Similarly, higher beta power was also associated with lower V_rms_ (β = -0.14 [-0.24 -0.050], t = 3.03, *p* = 0.0081). Beta burst duration was not significantly associated with V_rms_ (β = -0.089 [-0.18 0.0012], t = 1.94, *p* = 0.16).

**Figure 1.**
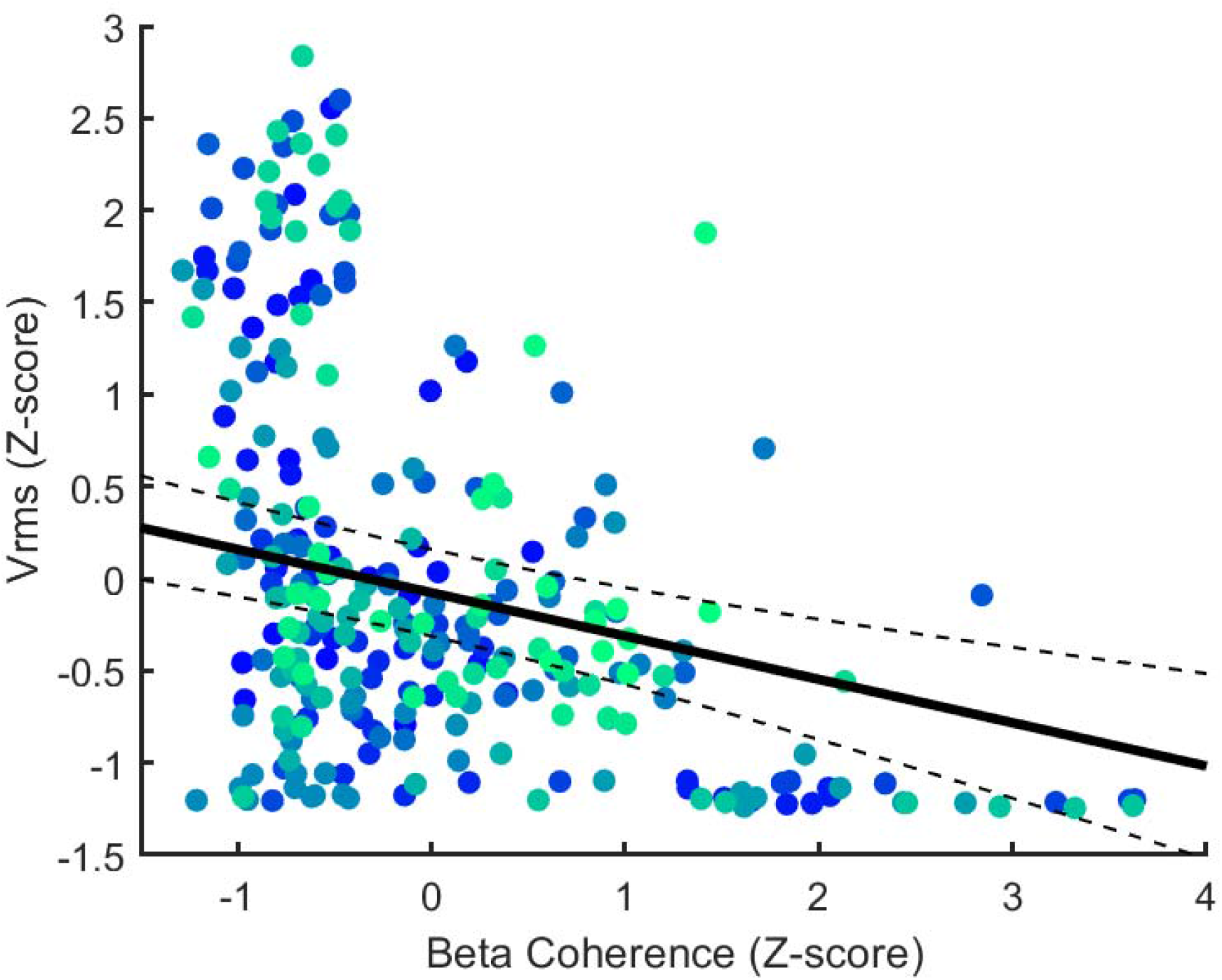
Higher Beta coherence is associated with worse bradykinesia. Scatter plot depicting beta coherence and V_rms_ for all participants across all visits. The gradient color depicts the participant ID. The solid line shows the average estimated slope of the relationship between beta coherence on V_rms_ and the dashed lines represent the 95% confidence interval for the linear mixed effects model.

### Bradykinesia Worsens Over Time

Figure 2A-C shows an example of the angular velocity trace during rWFE for one individual at three different time points after initial programming off therapy. At the three-month time point there was evidence of bradykinesia, as seen by the decrease in angular velocity and frequency of WFE cycles over the 30-second task, indicative of a sequence effect. At the twelve- and thirty-month time points, Fig. 2B and C respectively, bradykinesia was more severe, with a lower initial Vrms and more rapid deterioration in both the velocity and frequency domains. Visual comparison of the rWFE task at the different visits, Fig. 2A to C, demonstrated worsening of bradykinesia over thirty months in this individual. This was confirmed for the group overall, where a significant decrease (i.e., worsening) in V_rms_ was demonstrated over time (β = -0.015 [-0.022 -0.0073], t = 3.88, *p* = 1.31e-4; Fig. 2D).

**Figure 2.**
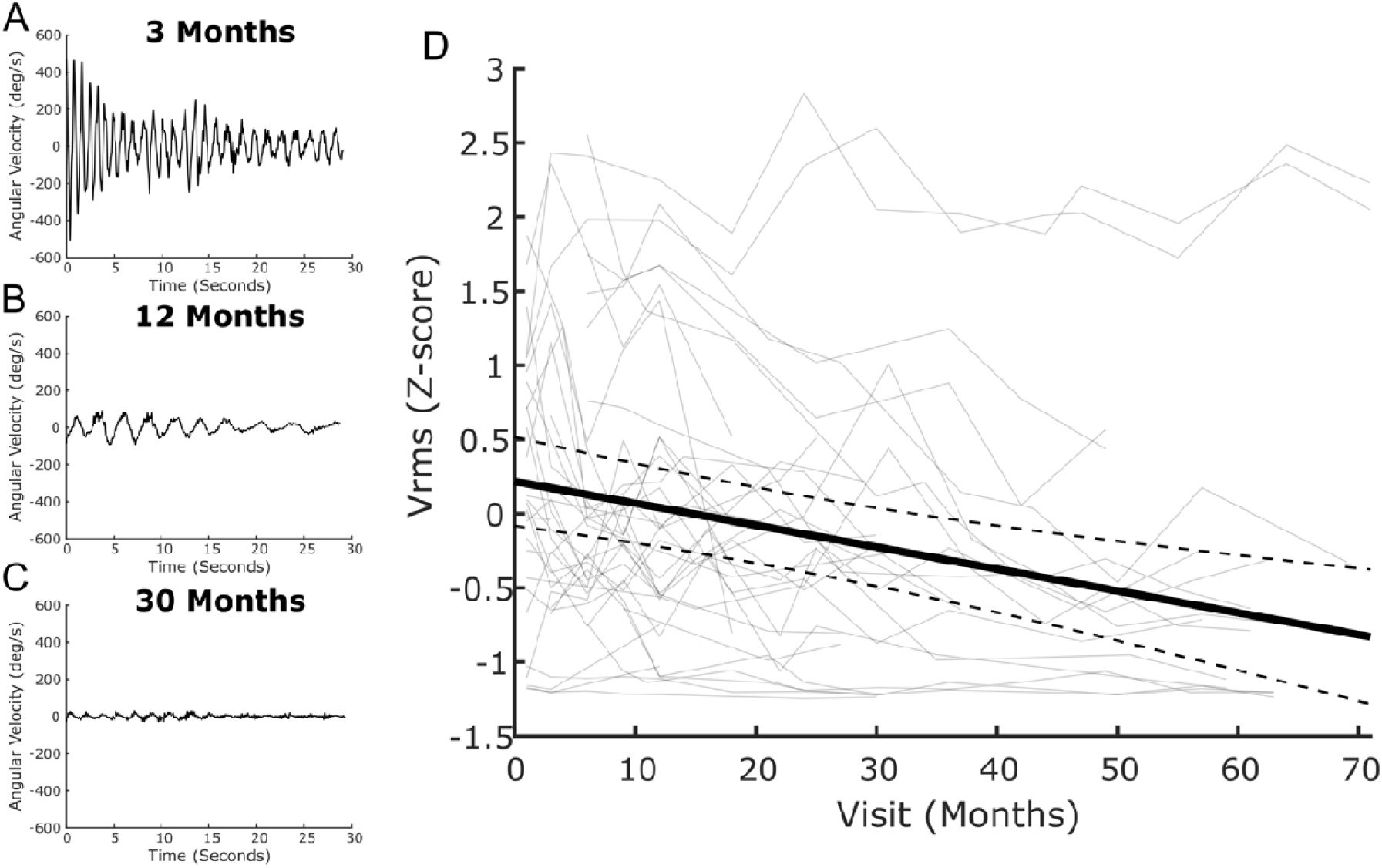
Bradykinesia worsens over time. (Left) Depiction of one example of angular velocity traces during rWFE at three different timepoints. The participant shows noticeable worsening of bradykinesia at later visits. (Right) Group results showing the effect of time on V_rms_. The gray lines depict individual’s performance at each visit. The solid line shows the average estimated slope of the effect of time on V_rms,_ and the dashed lines represent the 95% confidence interval for the linear mixed effects model.

### STN Beta Coherence Increases Over Time

Figure 3A-C shows an example of the observed coherence between STNs, off therapy, at three different time points after initial programming for one individual. The amount of coherence in the beta band, outlined by the dotted white line in the spectrogram, increased at each time point. This was confirmed when looking over the group, as a significant increase in beta coherence was observed over time (β = 0.0096 [0.0030 0.016], t = 2.85, *p* = 0.014; Fig. 3D). Meanwhile, neither normalized beta power (β = 0.011 [-0.00011 0.022], t = 1.95, *p* = 0.16) or beta burst durations (β = 0.0074 [-0.0035 0.018], t = 1.33, *p* = 0.54) significantly changed over time.

**Figure 3.**
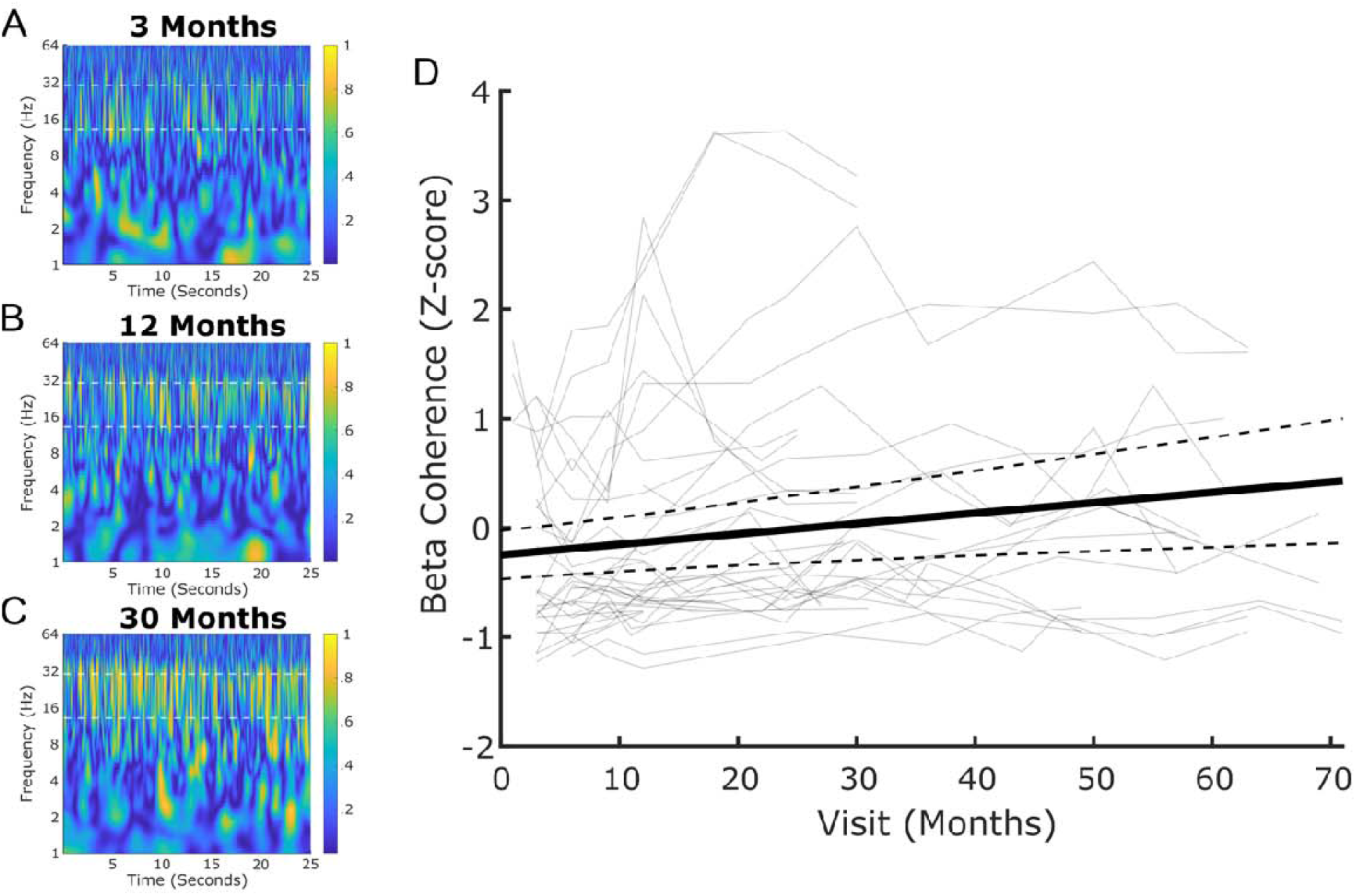
Beta Coherence increases over time. (Left) Depiction of one example of wavelet coherence during rWFE at three different timepoints. The dashed white lines denote the beta band (13-30 Hz). The color bar represents the magnitude squared coherence. The participant shows noticeable increase in coherence within the beta band as time passes. (Right) Group results showing the effect of time on beta coherence. The gray lines depict individual’s coherence at each visit. The solid line shows the average estimated slope of the effect of time on beta coherence and the dashed lines represent the 95% confidence interval for the linear mixed effects model.

### Changes in Bradykinesia are Related to Changes in STN Beta Coherence

Figure 4 shows examples from two different participants: one who experienced significant worsening of bradykinesia over time (Fig. 4A) and one who did not demonstrate worsening of bradykinesia over time (Fig. 4B). The patient who experienced progressive worsening of bradykinesia, off therapy, over the time course of thirty months after initial programming also demonstrated a progressive increase in STN beta coherence. Meanwhile, the participant who did not experience any substantial worsening of bradykinesia over time did not display an increase in STN beta coherence. When looking across the group, the percent change in beta coherence per month was significantly related to the percent change in V_rms_ per month (B = -0.96 [-1.59 -0.33], t = 3.01, *p* = 0.0087; Fig 4C). A negative association was observed where larger increases in beta coherence were associated with larger decreases (i.e., worsening) of V_rms_. Meanwhile, neither percent change in beta burst durations (B = -0.085 [-0.19 0.019], t = 1.60, *p* = 0.33) or normalized beta power (B = -0.070 [-0.16 0.021], t = 1.52, *p* = 0.39) significantly related to percent change in V_rms_.

**Figure 4.**
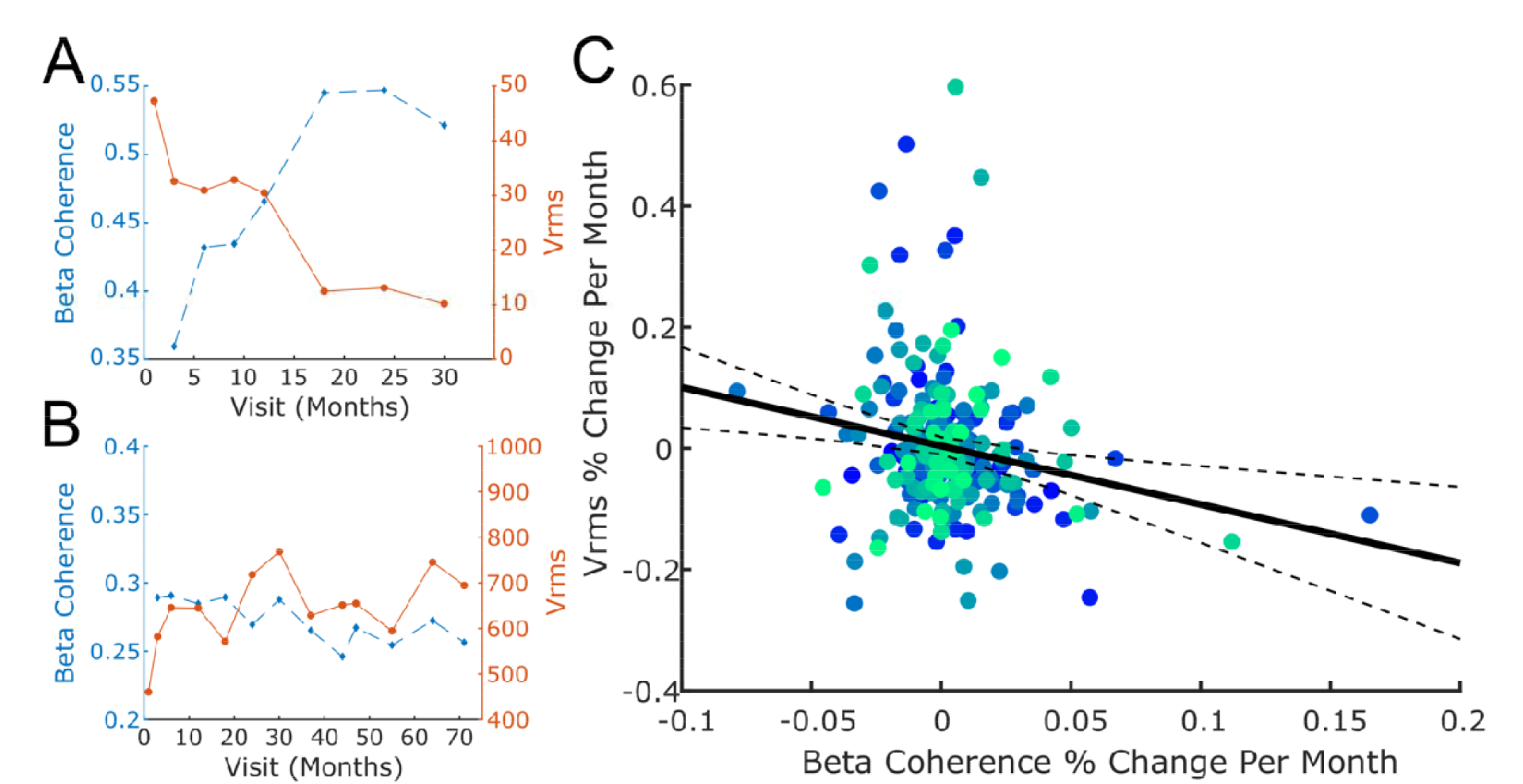
Changes in Beta Coherence Over Time are Related to Changes in Bradykinesia. (Left) Two different case examples of trajectories of beta coherence and V_rms_ over time. The top example shows an inverse relationship between beta coherence and V_rms_. There is a noticeable worsening of bradykinesia (i.e., decrease in V_rms_) that is mirrored by an increase in STN-STN beta coherence. Meanwhile, the bottom example shows an example of someone with minimal bradykinesia (V_rms_ above 600 deg/s) which is stable over time. Their beta coherence, similarly, does not increase over time. (Right) Scatter plot of individuals depicting the percent change per month for beta coherence compared to the percent change per month for V_rms_ for each of the visits. The gradient color depicts the participant ID. The solid line shows the average estimated slope of the relationship between percent change per month of beta coherence on the percent change per month of V_rms_ and the dashed lines represent the 95% confidence interval for the linear mixed effects model.

## Discussion

The current study combined synchronized subthalamic neural and kinematic recordings during a validated quantitative assessment of bradykinesia with individuals with PD who were implanted with a first generation sensing neurostimulator (Activa® PC+S, Medtronic PLC) as part of a longitudinal ‘washout’ therapy protocol. Individuals were followed for up to seven years, with visits every three to six months (average three years). The results of this longitudinal study demonstrate that there was a progressive worsening of bradykinesia over time, off therapy, after a washout of both STN DBS and medication, and that this was related to progressively increasing interhemispheric neural synchrony between STNs within the beta band. Meanwhile, beta band power and burst dynamics did not significantly increase over time. These results highlight the potential role of heightened interhemispheric STN beta synchrony as a contributor to the progressive worsening of bradykinesia in moderate to advanced idiopathic Parkinson’s disease.

### Worsening of Bradykinesia Over Time

The UPDRS III and MDS-UPDRS III represent the most common methods for following clinical motor progression of Parkinson’s disease. However, these clinical scales have well-documented limitations for tracking symptoms in PD due to their lack of resolution^32,33^. Here, we used a validated repetitive wrist flexion-extension task with a wearable sensor to provide high-resolution quantification of behavior to understand the progression of upper limb bradykinesia^29^. We combined this with a 12- to 48-hour washout of medication and 60–75 minute washout of STN DBS to provide longitudinal tracking of underlying disease pathology. It has been demonstrated that tremor stabilized two years after STN DBS, and we have confirmed this in our washout study^22–26^. In contrast these results demonstrate that bradykinesia continues to worsen over time off therapy following chronic STN DBS, supporting other reports^23,24^. We recently showed a similar worsening of performance of repetitive alternating finger tapping, using quantitative digitography (QDG) technology over time, off therapy^34^. Specifically, the metrics correlated with MDS-UPDRS III bradykinesia worsened over time, whereas those correlated with MDS-UPDRS III tremor did not. Castrioto and colleagues found that the UPDRS III assessment of bradykinesia assessed in the off-therapy state declined after 10 years of STN DBS^23^. No worsening of bradykinesia was observed at one or five years in their study, whereas we observed worsening in most cases within the first year, Fig. 2D. The lack of observed worsening in bradykinesia at five years in the Castrioto study may be due to a combination of the lower resolution of the UPDRS III compared to the rWFE task used in the present study, as well as a less stringent therapy washout protocol (i.e., only overnight withdrawal of dopaminergic medication). This highlights the benefit of using high-resolution quantification of behavior via wearables for better insight into disease pathology and progression as compared to relying solely on clinical scales.

### Heightened Interhemispheric Beta synchrony in the STN

In the current study, higher interhemispheric beta synchrony between STNs was associated with worse bradykinesia. Previous studies have established a beta oscillopathy within the STN and throughout the basal ganglia underlying aspects of impairment and bradykinesia in Parkinson’s disease^5,35,36^. This heightened beta synchrony within the basal ganglia appears to also spread to the motor cortex^12,37^. The results of this study point to pathological beta synchrony in the bilateral sensorimotor network. The existence of this bilateral network pathophysiology is further supported by the observed similarities in peak beta frequencies (e.g., beta profiles) between both STN and motor cortex, as well as between STNs^8–10,38,39^. Additionally, it builds on the observed finding showing the presence of interhemispheric STN coherence off therapy^21^ as well as increased overlap of beta bursts between STNs in the off-levodopa state^40^.

Previous work on heightened beta synchrony in PD has focused on the relationship between STN and the motor cortex due to the known presence of direct connections via the hyperdirect pathway, and therefore the likelihood that any abnormal synchrony within the basal ganglia could be propagating throughout the basal ganglia-cortical loop^9,11,41,42^. Meanwhile, there are no known direct anatomical connections between STNs. Therefore, the observed presence of synchrony between STNs observed here is likely through multi-synaptic connections, potentially via cortex and corpus callosum, cerebellar and thalamus synapses, or the brainstem. This fits within previous frameworks of bradykinesia that have pointed out potential cerebellar and cortical influences on bradykinesia in addition to disrupted basal ganglia circuitry^1^. The notion of an underlying bilateral oscillatory network is further supported by observed improvements in ipsilateral motor function, including bradykinesia, both transiently and long-term following unilateral STN DBS^16–20^. Electrophysiological evidence from unilateral STN DBS shows alterations in the discharge pattern of the contralateral STN, supporting a network connecting the two STNs^43,44^.

The major finding of this paper is that bilateral STN coherence continued to increase over time, and that the observed increase in STN coherence was associated with worsening of bradykinesia. To our knowledge, this is the first paper of its kind showing longitudinal multi-year subcortical recordings accompanied by high resolution kinematics and a prolonged medication and DBS washout to investigate the neural and behavioral correlates of progressive pathology in PD. We provide evidence that the worsening of bradykinesia over time, despite chronic STN DBS and medication, is related to heightened bilateral STN beta synchrony. In contrast to the observed increase in bilateral STN coherence over time, neither STN beta power or beta burst duration significantly changed over time, and neither were related to the observed worsening of bradykinesia. This replicates a previous finding from our lab where neither beta power or burst duration, as measured at rest, showed significant increases over three years^22^. The lack of increase in beta power within the STN implies that the observed increase in beta coherence over time is not a byproduct of changes in power, but rather evidence for a change in synchrony between the two STNs.

### Limitations

One potential limitation of the current study is the degree to which therapy was fully ‘washed out’ prior to testing. The current ‘washout’ protocol involved stopping long-acting dopaminergic medication 24-48 hours and short-acting dopaminergic medication at least 12 hours prior to testing and turning STN DBS off for 60-75 minutes prior to completion of the rWFE task. We have previously shown that the effects of STN DBS on STN LFPs are washed out well before the 60-75 minutes used here^28^. Bradykinesia has been shown to be substantially washed out prior to 60 minutes^45,46^ and 60-90 minutes^47^ after turning off DBS. Additionally, a slow washout of therapy likely would have masked the observed worsening of bradykinesia, rather than artificially inflate it. Another potential concern is the effect of volume conduction on synchrony measures such as coherence. We used a bipolar recording configuration for each lead to minimize this potential confound, which has successfully been used in the past^10,37,48^.

## Conclusion

The results of this study elucidate a neural correlate of bradykinesia through the largest longitudinal therapy washout study to date combining subcortical LFP data and kinematic data in Parkinson’s disease. Specifically, heightened beta synchrony between STNs was related to worse bradykinesia off therapy, and worsening of bradykinesia over the time course of several years was related to increases in STN beta synchrony implying the existence of a pathological bilateral network. The growing availability of commercial sensing neurostimulators and use of wearable sensors in monitoring Parkinson’s disease should allow for further understanding of the potential neural mechanisms underlying symptom-specific progression.

## Data Availability

All data produced in the present study are available upon reasonable request to the authors.

## Acknowledgements

We would like to thank the members of the Human Motor Control and Neuromodulation laboratory, Dr Jaimie Henderson, and, most importantly, the participants who dedicated their time to this study.

## Author Contributions

KBW, YMK, MNP, RWA, and HMBS contributed to the conception and design of the study. KBW, YMK, MNP, RWA, JEP, MHT, RSN, MMK, AV, ZB, EJQ, and HMBS contributed to the acquisition and analysis of the data. KBW, YMK, MNP, HMBS contributed to drafting the text and preparing the figures.

